# Working with the future for the future: a peer-led educational intervention on antimicrobial resistance; a quasi-experimental study

**DOI:** 10.64898/2026.07.04.26357267

**Authors:** Sagar Mani Pradhan, Aparna Chakravarty, Anandu Hari, Vrinda Nampoothiri, Kusum Rani, Fabia Edathadathil, Sanjeev Kumar Singh

## Abstract

Antimicrobial resistance (AMR) is a global threat to public health and development. Failure to address it could return society to a ‘pre-antibiotic era’ with increased morbidity and mortality. Because human behaviour is crucial to AMR management, interventions modifying knowledge, attitudes, and practices are therefore essential. Modifying health-related behaviours presents a significant challenge, yet it is crucial for public health. Engaging populations during periods of shifting perceptions can address this challenge and ensure the sustainability of interventions. Adolescents and young people attending school represent a key demographic. The primary objective of this study is to evaluate students’ awareness, perceptions, and behaviours concerning antimicrobial resistance (AMR) and hygiene, while enhancing and empowering children as agents of change within the community. In this study, students from Allied Health Sciences (AHS) across various disciplines were recruited to serve as peer educators for an evidence-informed educational workshop. A pilot delivery of these activities was conducted among a few students in a school before the final delivery was executed in three schools. Schools following a comparable educational board and curriculum were selected for inclusion in the study. A structured questionnaire was employed to assess the effects before and after the intervention. Statistically significant improvements were observed in participants’ knowledge, attitudes, and practices (p < 0.001). Additionally, feedback was collected from participants, teachers, and the school nurse attending the session. By triangulating these findings, a notable immediate improvement was observed in students’ knowledge, attitudes, and practices. This study provides evidence that employing multimodal teaching led by peer education is a valid and effective method for delivering health messages. It further underscores the mutual benefits for stakeholders (peer educators and peer learners) by offering a two-way learning opportunity. The benefits extend beyond academic and core scientific learning to include increased confidence as effective health educators and future-ready healthcare professionals.

Fig 1.
Graphical representation of the study

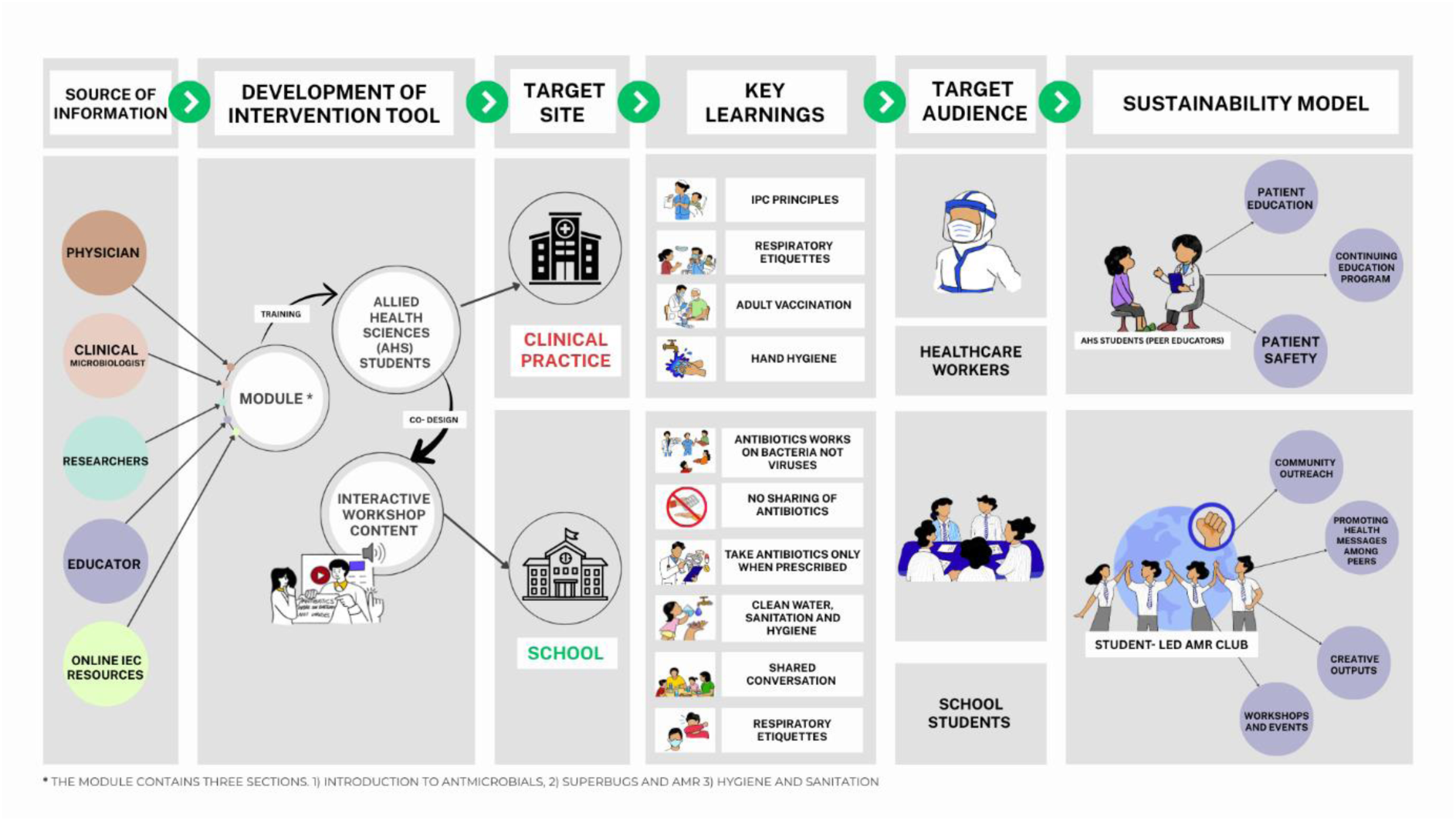

## 1. Introduction

Antimicrobial agents are employed to eliminate or inhibit the proliferation of microorganisms, including bacteria, viruses, fungi, and protozoa. Antimicrobial resistance (AMR) arises when microorganisms develop resistance, a process expedited by human activities, particularly the misuse and overuse of medications. Consequently, when these agents lose efficacy, infections become challenging to treat, thereby elevating the risks of disease transmission, disability, and mortality [1]. AMR represents a significant global public health threat. In 2021, bacterial AMR was responsible for an estimated 4.71 million deaths, with projections indicating 39.1 million deaths from 2025 to 2050, equating to three deaths per minute [2]. In India, 2021 saw an estimated 267,000 (224,000-310,000) deaths directly attributable to AMR and 987,000 (855,000-1,120,000) associated with it. These figures surpassed deaths from neoplasms, transport injuries, diabetes and kidney diseases in the same year [3]. Inappropriate, excessive antibiotic use significantly contributes to the development and dissemination of antimicrobial resistance (AMR). Self-medication and the availability of antimicrobials without a prescription further intensify this issue. Notably, India has experienced a twofold increase in antibiotic consumption, making it one of the largest consumers of antibiotics worldwide. [4, 5]

AMR is a multi-sectoral issue, and failure to address it may lead to a regression to a ‘pre-antibiotic era,’ causing significant morbidity and mortality. The severity of the issue is now being acknowledged, prompting action from global health organizations and governments [6]. Without preventive measures, AMR could result in 10 million annual deaths globally by 2050 [7]. In 2015, the World Health Assembly adopted the Global Action Plan on AMR (GAP-AMR), urging member states to formulate local national action plans (NAP). India’s NAP-AMR, introduced in 2017, employed an integrated One Health approach, outlining six strategic objectives under a ‘One Health Approach.’

Despite these efforts, they may not effectively resonate with the public. Therefore, a more straightforward, comprehensive narrative is necessary to convey the seriousness of AMR to the population [8]. The updated GAP-AMR identifies “strengthen awareness and promote appropriate social and behavioural change to reduce antimicrobial resistance risks across all sectors” as its first strategic objective [9]. The 2016 review on AMR, chaired by Jim O’Neill, is a globally recognized report that serves as a wake-up call. Among the nine recommended interventions, a global public awareness campaign was prioritized [10]. India’s NAP-AMR also emphasizes improving awareness as the foremost of its six strategic priorities [11]. Prioritizing effective communication as the first step is not merely incidental placement but a crucial activity for addressing AMR within the community.

Public health initiatives have proven effective in enhancing understanding and modifying behaviours [12, 13, 14, 15]. Understanding human behaviour facilitates the development of effective communication strategies. However, a significant challenge lies in the societal framework, which comprises rational, irrational, and rationalizing individuals. Consequently, a universal model cannot adequately address this complex structure to achieve desired outcomes. The most effective strategy involves segmenting the audience and tailoring interventions accordingly.

Modifying health-related behaviours is a considerable challenge, yet it is vital for public health [16]. Addressing populations during periods of changing perceptions can address this issue and ensure the sustainability of interventions. Adolescents and young people attending school constitute a key demographic. According to WHO, adolescents are aged 10-19 years, while youth are 15–24 years [17]. During these formative years, they are receptive to adopting health behaviours and can influence change within families, communities, and healthcare settings. This group encompasses diverse demographics: AMR survivors, indigenous voices, marginalized and privileged communities, antimicrobial users, laypersons, professionals, and future generations [18]. Engaging with young people helps bridge awareness gaps and fosters a generation more capable of responsible antimicrobial use.

Our study aimed to assess students’ awareness, perceptions, and behaviours concerning AMR and hygiene, empowering children as agents of change. We involved school children as peer learners and healthcare students as peer educators, establishing peer education as a fundamental component for enduring behaviour change.

By providing children with information on AMR, infection prevention, and the prudent use of antibiotics, and by training healthcare students to convey these messages engagingly, the study sought to achieve two objectives: increasing awareness among students and enhancing communication and leadership skills, while promoting antimicrobial stewardship among future professionals. This strategy addresses AMR as a public health issue by cultivating informed citizens and accountable healthcare workers who can collaborate to mitigate the spread of antimicrobial resistance.

## 2. Materials and methods

### 2.1. Ethical consideration

The study was conducted as part of S.M.P.’s (first author on this study) fellowship program, for which ethical approval for the entire study was obtained from the Institutional Ethics Committee, Amrita Hospital, Faridabad, Haryana (AIMS-IEC-BAS-09-25-003). Written informed consent was obtained from the participant and their parents before conducting the session. To address low participation and acceptance, incentivised participation was used as a risk mitigation strategy.

### 2.2. Study design

The study employed a single-arm, school-based, quasi-experimental design, conducted without a control group, across four schools in Delhi and the Delhi-NCR region. It involved 257 students in the final study and 30 in the pilot phase, all of whom met the inclusion criteria outlined in Table 1. Workshops were held during regular school hours and on school premises to ensure accessibility and minimize disruption. These workshops were collaboratively facilitated by allied healthcare students and the author, utilizing a peer-to-peer teaching approach. Data collection involved administering pre- and post-workshop assessments to all student participants, with participation in the assessments being voluntary.

**Table 1.** Selection criteria for the study.

|  | INCLUSION | EXCLUSION |
| --- | --- | --- |
| STUDY SITE | Schools that follow the National Council of Educational Research and Training (NCERT) Curriculum under the Government of India.<br>Schools that operate under the Central Board of Secondary Education (CBSE), Government of India board. | Schools that follow any curriculum other than NCERT.<br>Schools affiliated with boards other than CBSE. |
| PARTICIPANTS | • Students enrolled in standard ninth and tenth.<br>• Provide consent and assent from parent/ guardian. | • Students enrolled below standard ninth and above standard tenth.<br>• The student is unwilling to participate or not available during the workshop. |

### 2.3. Selection of participants and site

The schools selected for participation were affiliated with the same educational board and adhered to an identical curriculum (Table 1). This strategy minimised variability due to differences in curricula and educational policies, thereby enhancing the study’s internal validity. The participant selection process employed a strategic, criteria-based methodology. The study population comprised secondary school students, who are at a crucial developmental stage where behavioural changes can be established and often persist into adulthood (Table 1).

Thirteen allied health sciences students representing diverse fields including radiology and imaging technology, emergency medicine technology, cardiovascular technology were recruited for peer educators based on their proficient communication skills with peers, their role as relatable models, and their capacity to facilitate the continuous dissemination of health information in clinical settings. This approach enhances engagement, acceptability, and potential behaviour change compared to traditional educator-led methods, while also supporting the intervention’s long-term sustainability.

Approval was secured from the head of the allied health sciences institute prior to the study. Interested student volunteers were provided with a brief overview of the study. Those who consented to participate underwent a training program, mentored by clinicians and researchers.

### 2.4. Workshop structure

A multimodal teaching model was employed for all workshops. This approach aimed to enhance students’ conceptual understanding of the topic, facilitate the application of knowledge to real-world contexts, and raise awareness of the topic’s significance and urgency. An initial pilot study was conducted in one school before the main study. This pilot served as a pre-validation for the final workshops. Based on feedback from students and faculty who attended the pilot, the following improvements were implemented:

1. Workshop Duration: The pilot helped determine the appropriate duration to maintain participant engagement throughout the workshop.
2. Reorganisation of Activities: Based on observations and outcomes, certain activities were refined. The pilot also identified areas where students tended to lose focus. Integrating enjoyable activities and games in these areas helped maintain attention and active participation.

The workshop was conducted over two days at each school, delivered to small groups of 40-50 students each day by 6-7 peer educators to enhance understanding and comprehension. Peer educators were divided into two batches of 6-7 students each, visiting the study site alternately. This arrangement minimized disruption to their academic and clinical responsibilities. The workshop concluded with students and the faculty voluntarily completing a feedback form.

Each workshop session lasted three hours and included an interactive teaching module using Microsoft PowerPoint for Microsoft 365 MSO (Version 2601 Build 16.0.19628.20132). The workshop included room for discussions, a query-solving session, games, an art reflection worksheet (S4. Fig. 3), an audio-visual display, and role-plays (S5.). To increase adherence, incentives such as themed caps for students and illustrative books for the school were provided.

### 2.5. Development of educational module and workshop

The curriculum covered topics including antimicrobial resistance, antibiotics, and hygiene practices. The module’s content was developed through consultations with clinicians, researchers, and educators. Various online information, education, and communication (IEC) materials were incorporated into the module. (S1.Table 1).

The student volunteers of allied health sciences participated in a pre-assessment, followed by comprehensive training on antimicrobial resistance (AMR) and behavioural conduct, informed by the experiences of clinicians and researchers. This training concluded with a post-assessment of the volunteers. Subsequently, these volunteers played an active role in co-designing the interactive components (audio-visual aids, demonstrations, hands-on activities) of the workshop.

### 2.6. Data collection instrument

A structured questionnaire was developed through consultations with clinicians and researchers at Amrita Hospital, Faridabad, as well as an educator in Delhi NCR, to ensure content validity and context-specific relevance (S2. Fig. 1) The questionnaire consisted of four primary sections: a) informed consent, b) anonymized identification code and socio-demographic details, and c) a knowledge, attitude, and practices (KAP) section, which included five statements each related to antibiotic use, antibiotic resistance, hygiene practices, and the One Health concept.

Utilizing a 3-point Likert scale, the options of agree, uncertain, and disagree were included across three domains, each comprising five statements. Each correct or appropriate response was awarded 2 points, while incorrect or inappropriate responses received no points, and uncertain responses were given 1 point. An overall score exceeding 80% was considered indicative of good KAP.

### 2.7. Data analysis

The data collection involved a structured questionnaire administered both before and after the intervention, with responses self-reported by participants. The collected data were initially entered into Microsoft® Excel® for Microsoft 365 MSO (Version 2601 Build 16.0.19628.20214) 64-bit for processing and subsequently exported to Python (Version 3.10.9) for statistical analysis. A psychometric analysis was conducted to assess the construct validity and reliability of the survey instrument. Baseline responses for each questionnaire item were summarized as frequencies and percentages (n, %), and the distributions were examined for potential ceiling effects. Internal consistency was evaluated using Cronbach’s alpha coefficient. Domain scores from pre-intervention and post-intervention were compared through paired analysis. The normality of change scores (post minus pre) was assessed using the Shapiro-Wilk test, and the Wilcoxon signed-rank test was applied due to observed deviations from normality. Despite employing non-parametric testing for conservative statistical inference, mean ± standard deviation (SD) values were reported to aid in interpreting the magnitude of change. The intervention effect size was quantified using Cohen’s d.

In addition to the analysis of continuous scores, domain scores were categorized using a modified Bloom’s cut-off (≥80% of the maximum score). McNemar’s test for paired binary data was utilized to evaluate changes in the proportion of pre- and post-scores.

Content analysis was performed on feedback forms completed by both students and faculty attendees. The findings were triangulated with quantitative results using MS Excel to establish convergent validity.

## 3. Results

### 3.1. Background characteristics

The final study was conducted among 257 students enrolled in the ninth and tenth grades, respectively at the time of the study. The study was conducted at three school sites in Delhi and Faridabad (Delhi NCR region). The cohort included 144 male students (56.03%) and 113 female students (43.96%) within the range of 13-17 years of age.

### 3.2. Knowledge, attitude, practice (KAP) findings

Only three participants (1.1%) reported to have been exposed to an antibiotic awareness campaign prior to this study (Table 2). This might explain the baseline assessment results with only 6.6% students to have good knowledge. A substantial knowledge gaps were observed in the understanding of antibiotic use, where more than half of the participants incorrectly agreed that antibiotics have the potency to kill viruses (61.1%) and cure most of cough and cold (68.1%) (Fig. 2). The students, however, relatively scored correct responses for statements on unnecessary use of antibiotics making it ineffective (70.8%) and antibiotic resistance being a problem in the country and globally (65.4%), suggesting partial prior awareness (Fig. 2). The majority of the students were either incorrect (51.0%) or uncertain (38.9%) of antibiotic use among animals reducing the effect of antibiotics in humans, reflecting limited understanding of ‘One Health Approach’.

**Fig 2.**
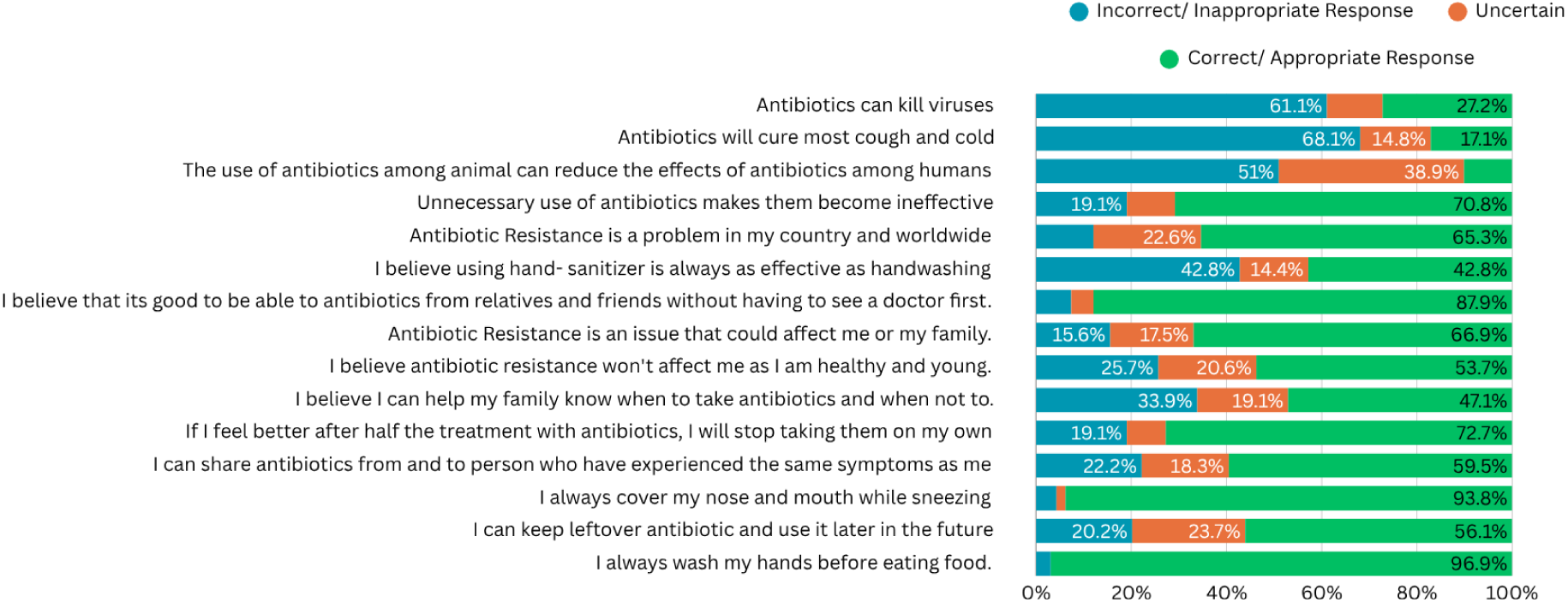
Baseline distribution of responses before intervention (n=257)

**Table 2.** Socio-demographic details of the participants.

| Variable | Baseline Responses |  |
| --- | --- | --- |
|  | Frequency (n) | Percentage (%) |
| Gender |  |  |
| Male | 144 | 56 |
| Female | 113 | 44 |
| All | 257 | 100 |
| Age |  |  |
| 13- 14 (Young people) | 142 | 55 |
| 15 -17 (Youth) | 115 | 45 |
| All | 257 | 100 |
| Have you ever been exposed to an antibiotic campaign before? |  |  |
| No | 254 | 99 |
| Yes | 3 | 1 |
| Total | 257 | 100 |

A majority of students (87.0%) believed that one should get antibiotics on doctors’ advice and not from relatives or friends (Fig. 2). More than half of the participants (66.9%) acknowledged that antibiotic resistance could affect them or their family, whereas 53.7% agreed that it could affect them regardless of being healthy and young. For the item stating, “if the students can help their family know when and when not to take antibiotics”, 33.9% disagreed while 19.1% were uncertain. The proportion of students who believed hand sanitizer is always as effective as handwashing (42.8%) was equal to those who did not believe this (42.8%), with a small proportion unsure (14.4%) about it.

The practice items demonstrated predominantly appropriate responses. Most students (72.8%) agreed on not discontinuing antibiotics on their own once they start feeling better. Over half of the study cohort (59.5 %) disagreed on sharing antibiotics despite having similar symptoms (Fig. 2). A large majority of the students responded appropriately for hygiene practices-always covering nose and mouth when sneezing (93.8%) and always washing hands before eating food (96.9%) (Fig. 2).

### 3.3. Impact of the intervention on the school students

The knowledge scores increased significantly following the intervention (p<0.001), with a mean improvement of 1.86 points and a large effect size (d= 0.846) (Fig. 2), indicating substantial educational impact. The participants’ post-intervention exhibited better understanding of antibiotics, wherein 66.9% correctly disagreed to item list, “antibiotics can kill viruses” and 34.6% disagreeing on “antibiotic will cure most cough and cold” (Fig. 2). Understanding of one health in the item list “the use of antibiotics among animal can reduce the effects of antibiotic among humans” shows more participants (36.6%) agreeing to it (Fig. 3).

**Fig 3.**
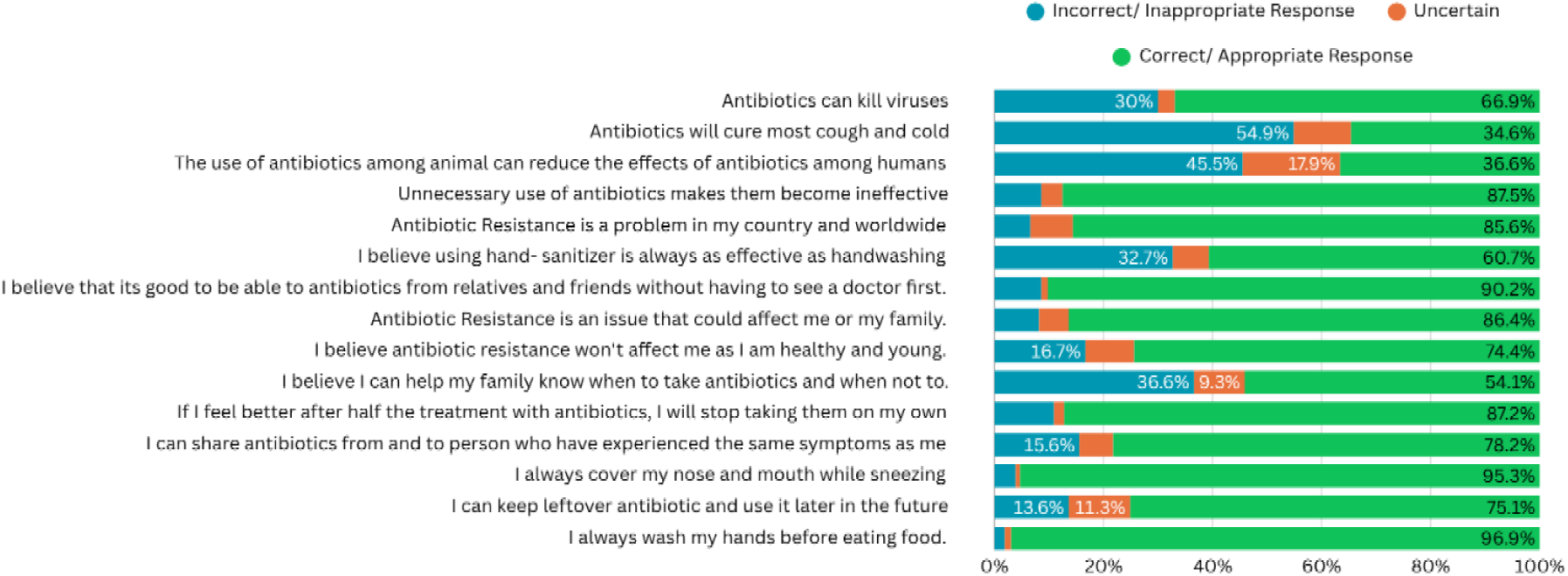
Baseline distribution of responses after the intervention (n=257)

Attitude scores also demonstrated significant improvement (p < 0.001), with a moderate effect size (d = 0.425), reflecting meaningful positive shifts in perception toward antibiotic resistance. Across all the item lists, their role in helping family know when and when not to take antibiotics showed comparatively smaller improvements (54.1%). Higher students (60.7%) post-intervention correctly disagreed on item list “hand sanitizer is always as effective as hand-washing” (Fig. 3). A positive change in perception of antibiotic resistance being an issue that could affect anyone was observed as substantial proportion of the students rightly agreed to it (86.4%) (Fig. 3).

The predominance of correct response was clearly observed in the baseline assessment for the practice domain (Fig. 2). The highest correct response recorded was for item lists on hygiene practices-always covering the nose and mouth while sneezing (93.8%) and always washing hands before eating food (96.9%) which persisted through to the post-intervention (Fig. 2, Fig. 3).

The proportion of students achieving good scores increased significantly across all domains (Table 3). To provide better interpretable estimates of improvement, domain scores were categorized using modified Bloom’s cut-off (≥80% of maximum score). The proportion of students with good knowledge increased from 6.6% at baseline to 39.7% post-intervention (p < 0.001).Similarly, good attitude increased from 41.6% to 65.8%, and good practice increased from 69.3% to 85.2% (Table 4).

**Table 3.**
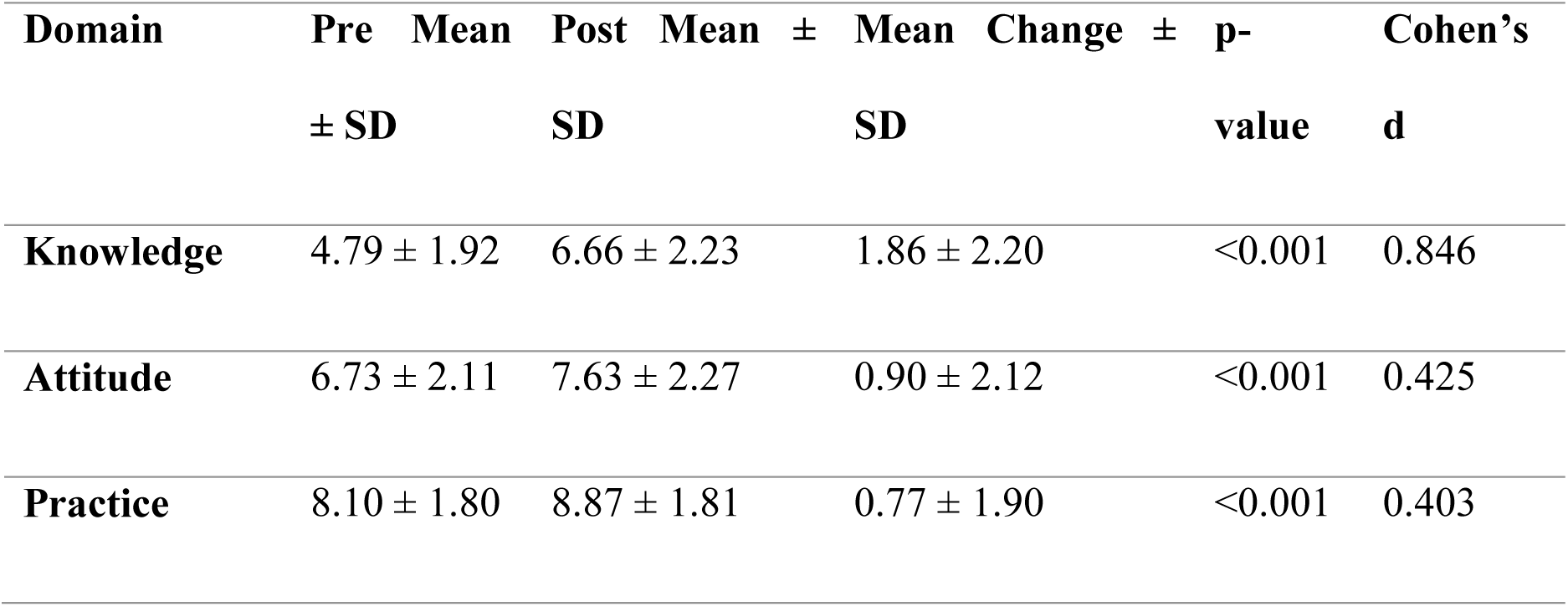
Pre-Post Comparison of domain scores (n=257)

**Table 4.** Change in proportion of students with good scores according to modified Bloom’s cut-off (≥80%) (n=257)

| Domain | Pre-intervention<br>Good n (%) | Post-intervention<br>Good n (%) | McNemar p-value |
| --- | --- | --- | --- |
| Knowledge | 17 (6.6%) | 102 (39.7%) | <0.001 |
| Attitude | 107 (41.6%) | 169 (65.8%) | <0.001 |
| Practice | 178 (69.3%) | 219 (85.2%) | <0.001 |

The improvements were evident at multiple levels of analysis. Item-level distributions demonstrated increased proportions of correct or appropriate responses across most knowledge and attitude questions following the intervention. Domain-level score comparisons confirmed significant increases in aggregate scores, with the largest magnitude of change observed in the Knowledge domain.

Categorical analysis using modified Bloom’s cut-off further demonstrated meaningful improvement in the proportion of students achieving adequate performance, particularly in knowledge and attitude domains (Table 4). While practice scores also improved, higher baseline performance and identified ceiling effects in selected items may have limited the observable magnitude of change.

Collectively, these findings indicate that the intervention was effective in improving awareness and perceptions related to antimicrobial resistance, with the most pronounced impact on knowledge acquisition. The comparison of knowledge, attitude and practices (KAP) scores pre intervention with post intervention highlighting the improvements is illustrated in Table 3.

**Fig 4.**
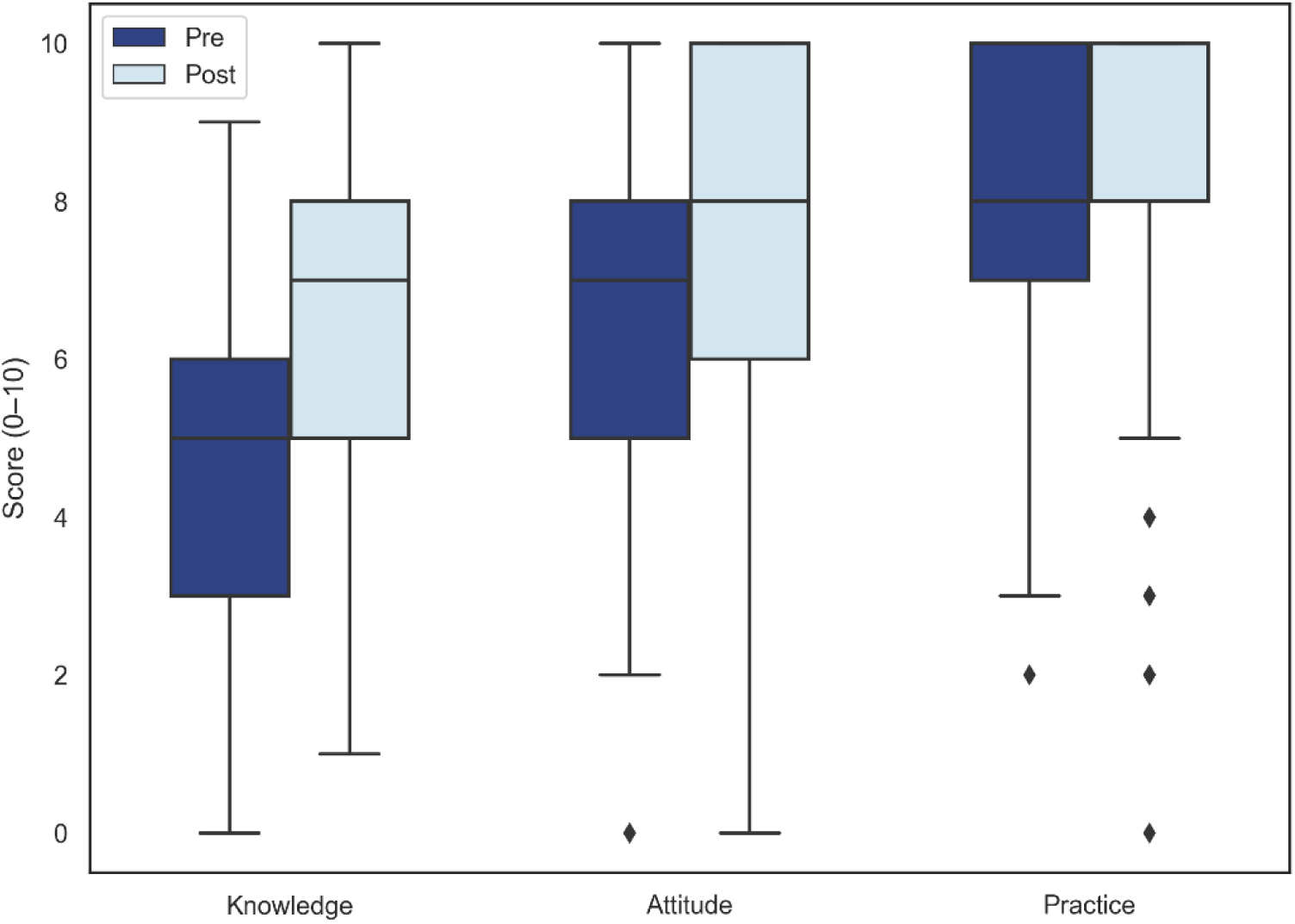
Comparison for pre and post KAP scores of the participants.

### 3.4. Feedback content analysis

For the purpose of triangulation of understanding the intervention impact, non-structured open-ended feedback was obtained at the end of each session from faculty staff (n=10) and consenting school students (n= 200). After the completion of the entire study across all the sites, 13 peer educators were also asked to fill in the form.

#### 3.4.1. Feedback from the students

Most of the students initially had a limited knowledge on antibiotics and antimicrobial resistance. The students post intervention articulated the learnings in the form using their own words suggesting cognitive and perceptual transformation as observed in Table 4.

> *“It can shake your financial condition because it is antimicrobial resistance.”-Student, School A*
>
> *“Antibiotics + Anti - advice = Antibiotic resistance.”-Student, School B*
>
> *“I will develop healthy habits and take part in building a healthier future for all.”-Student, School C*

Initially, most participants were unsure or did not agree that they could help their families know when and when not to take antibiotics. This might reflect poor involvement and sense of responsibility. Following the intervention, empowerment and behavioural intent seemed to have improved as the affirmations from the students encompassed antimicrobial stewardship commitment, awareness promotion and eagerness for future involvement positioning participants as motivated advocates and sustained impact.

> *“I shall now help my family and relatives by educating them about antibiotics.” - Student, School C*
>
> *“And now I have become a responsible health ambassador at my home and the community.” - Student, School A*

The module was designed primarily in a fashion that promotes engagement and adherence by the students. Audio-visual aids, demonstrations, hands-on activities were employed in a themed presentation. The positive feedback regarding this underscores how interactive approaches aided with emotional support and warm interaction from the facilitators boosts engagement and curiosity.

> *“It wasn’t a simple lecture, but a fun learning session filled with interesting activities. I liked the way it was superhero themed.” - Student, School A*.
>
> *“Had a perfect session with the respected team. They were well-trained and confident. The facilitators were well-dressed and attentive to our needs throughout the whole session. Overall, the session was great.”-Student, School C*

#### 3.4.2. Feedback from the faculty staff

The faculty staff from the selected schools including science teachers and school nurse were invited to attend the session. Feedback was collected from the faculty at the end of the session. This approach provides more complete and multi-perspective picture complementing the student’s feedback and strengthening the validity and practical utility of the study. The faculty staff provided their opinions on pedagogical goals, conceptual depth, and alignment with curriculum, bringing in a different lens, thereby triangulating the observation of whether the proposed objectives were met.

> *“The use of real-life examples, interactive discussions and visual aids helped simplify complex scientific concepts.”-Faculty Staff, School A*
>
> *The teacher/ resource person facilitated the session with clarity, enthusiasm, and professionalism.”-Faculty Staff, School B*
>
> *“Students showed a high level of interest and engagement throughout the session.”-Faculty Staff, School C*

The faculty reported on “learning something new” from the student-focused session. It reflects gains on new insights on daily practices contributing to AMR, affirming its educational value for school education and proving to be pedagogically active and professionally transformative as well.

> *“As a faculty, I too learned a lot about Antimicrobial Resistance and the importance of wise usage of antibiotics.”-Faculty, School C*
>
> *“The presence of Triclosan and Triclocarban in the soap and its relation to AMR was something I didn’t care for. I shall be careful from this moment onwards”-Faculty, School B*

The faculty reported gaps persisting in the current curriculum lacking information on AMR and displayed strong support for embedding AMR topics into the academic curricula. The faculty agreed that incorporating the topic into the school curriculum helps in building a healthier and conscious generation.

> *“Introducing AMR into the CBSE curriculum is a forward-looking step that aligns with India’s commitment to global health goals.”-Faculty, School A*
>
> *“Educating students on AMR creates awareness, encourages responsible behaviour and prepares them to face real-world health challenges wisely.”-Faculty, School A*

#### 3.4.3. Feedback from the facilitators

At the end of all the sessions, the allied healthcare students (peer educators), provided feedback. They reported that working as a facilitator in the session helped with improvements in their soft skills (communication, teamwork, leadership, socialising, etc.). Healthcare students who assumed the role of small group leaders reported that this experience facilitated the consolidation of their own knowledge. It also reinforced antimicrobial stewardship principles in their clinical practice and enhanced their roles as informed healthcare professionals, thereby fostering their development as effective science communicators.

> *“This experience also helped me improve my communication skills, reduce my stage fear and become more comfortable socialising and interacting with different people.”-Student, Program A*
>
> *“This (study) improved my communication skills and gave me practical exposure outside the classroom, helping me connect my theoretical knowledge with the real-world practice. My biggest personal achievement was gaining confidence.”-Student, Program C*
>
> *“Ever since the study, I have applied the lessons learnt to both my clinical and personal life. Adhering to correct techniques of handwashing, mindful use of antibiotics and sharing the same to my peers have been a part of my habit now.”-Student, Program E*
>
> *“It made me realise the important role a healthcare student can have in promoting awareness and supporting public health initiatives.”-Student, Program B*

## 4. Discussion

This school-based quasi-experimental study evaluated the immediate effect of an interactive antimicrobial resistance (AMR), hygiene, and One Health educational intervention among secondary-school students, delivered with the support of trained healthcare/allied-health student facilitators. The findings indicate that a structured, age-appropriate, activity-based educational module can improve students’ awareness and understanding of AMR-related concepts, responsible antimicrobial use, infection prevention, and hygiene practices. The use of peer-assisted facilitation and interactive learning methods such as demonstrations, games, role-play, and audio-visual content appeared to support engagement and comprehension among adolescents.

The improvement observed after the intervention is important because AMR is increasingly recognized as one of the most serious global public-health and development threats. It was also noted that only 1.1% of the students had prior exposure to AMR awareness campaign, thereby highlighting the gap in awareness among young adults. The present study contributes to this priority by demonstrating a practical school-based model for AMR communication among adolescents.

The intervention aligns with the first objective of the World Health Organization Global Action Plan on AMR, which emphasizes improving awareness and understanding through effective communication, education, and training [19]. It also supports the priorities of India’s National Action Plan on AMR, where awareness generation and community engagement are central components of a One Health response [20]. Importantly, the study did not restrict AMR education to antibiotic use alone, but also incorporated hygiene, infection prevention, and One Health concepts. This broader approach is relevant because AMR is driven not only by misuse and overuse of antimicrobials but also by poor infection prevention, inadequate hygiene, and environmental and animal-health interfaces [1].

The choice of school-going adolescents as the target group is a strength of the intervention. A meaningful positive change in attitude was observed among the participants, indicating that the intervention successfully shifted their perception in a constructive direction. Attitude change is a key predictor of behavioural change which makes this shift particularly important. This positive attitude change aligns with established theories of behaviour modification, which emphasize that cognitive and emotional engagement precedes sustained action. The results also reinforce the importance of selecting participants who already recognize the seriousness of an issue, as such individuals are more primed for meaningful change and less resistant to new perspectives.

Adolescence is a formative period during which health-related knowledge, attitudes, and habits are still developing. Educational interventions delivered during this stage may therefore influence not only individual understanding but also household-level conversations around antibiotic use, hygiene, and infection prevention. School students may act as information carriers within families and communities, particularly when educational content is simple, memorable, and action-oriented. Previous school-based AMR education initiatives, including e-Bug-based interventions, have shown that structured teaching resources can improve students’ knowledge of microbes, infection transmission, hygiene, and prudent antibiotic use [21, 22]. The present study adds to this literature by applying an interactive, superhero-themed, facilitator-led model in Delhi and the Delhi-NCR school context.

Another important feature of this study is the involvement of healthcare/allied-health students as peer educators. This created a dual educational pathway: school students received AMR-related education, while healthcare students gained experience in science communication, community engagement, and public-health leadership. Such models may be particularly useful in low- and middle-income settings, where scalable public-health education requires trained human resources and context-sensitive communication. The use of healthcare students as peer educators can also help bridge the gap between technical AMR knowledge and lay public understanding. In our study, the peer educators were trained to communicate the messages to the school students, thereby actively addressing the integration of antimicrobial stewardship principles among these volunteers, which would later be implemented into their clinical practice, further strengthening the future to combat AMR.

The content feedback further supports the acceptability of the intervention. Students’ and teachers’ responses suggested that interactive delivery, visual materials, demonstrations, and relatable examples helped make AMR understandable. This is important because AMR is often perceived as an abstract scientific issue rather than a problem linked to everyday behaviours such as incomplete antibiotic courses, self-medication, poor hand hygiene, and unnecessary antibiotic use for viral illnesses. By translating AMR into practical messages, the intervention may have improved perceived relevance and reduced communication barriers.

The findings should be interpreted in light of certain limitations. First, the study used a single-arm pre–post design without a control group; therefore, improvements cannot be attributed exclusively to the intervention with the same confidence as in a randomized controlled design. Second, the post-assessment was conducted immediately after the workshop, so the results primarily reflect short-term improvement in knowledge and attitudes rather than long-term retention or sustained behaviour change. Third, responses were self-reported and may have been influenced by social desirability bias, especially for hygiene and antibiotic-use practices. Poor demonstration of handwashing technique and respiratory etiquette by the students during activity supports this argument. Fourth, the study was conducted in selected schools in Delhi and the Delhi-NCR region, which may limit generalizability to rural settings, other states, or different socio-cultural contexts. Finally, facilitator-related variability may have influenced delivery despite prior training.

Despite these limitations, the study provides evidence that school-based, interactive AMR education is feasible, acceptable, and potentially effective for improving short-term awareness among adolescents. Future studies should include a control group, longer follow-up assessments, larger and more diverse school populations, and objective or behaviour-linked indicators where feasible. A cluster-randomized or stepped-wedge design could provide stronger evidence of effectiveness. Integration of AMR education into school health programs, science curricula, and community outreach activities may help sustain awareness and contribute to India’s broader One Health AMR response.

## 5. Conclusion

This study demonstrates that an interactive, school-based educational intervention on antimicrobial resistance, hygiene, infection prevention, and One Health can improve awareness and understanding among secondary-school students. The use of trained healthcare/allied-health student peer educators, activity-based learning, role-play, demonstrations, and age-appropriate communication made AMR more accessible to adolescents and created a dual benefit by strengthening public-health communication skills among future healthcare professionals.

The findings support the inclusion of AMR awareness activities in school health education and community engagement programs. Future research should include controlled designs, longer follow-up, larger and more diverse student populations, and assessment of sustained behavioural outcomes. Scaling such interventions through schools may contribute meaningfully to national and global AMR action by building a generation that understands responsible antimicrobial use, hygiene, and infection prevention.

## Data Availability

Due to ethical and privacy considerations, the datasets generated and/or analyzed during the current study are not publicly available. Researchers with a reasonable request may obtain access to the data by contacting the corresponding author at the email address provided in the manuscript, subject to approval by the relevant ethics committee and in accordance with applicable institutional and participant confidentiality requirements.

## Author contributions

Conceptualization: Sagar Mani Pradhan, Aparna Chakravarty, Kusum Rani, Sanjeev Kumar Singh

Data Curation: Sagar Mani Pradhan, Anandu Hari, Vrinda Nampoothiri

Formal analysis: Anandu Hari, Sagar Mani Pradhan, Vrinda Nampoothiri

Investigation: Sagar Mani Pradhan, Anandu Hari

Methodology: Sagar Mani Pradhan, Aparna Chakravarty, Sanjeev Kumar Singh, Kusum Rani, Vrinda Nampoothiri

Project administration: Sagar Mani Pradhan, Aparna Chakravarty, Fabia Edathadathil

Resources: Sanjeev Kumar Singh, Aparna Chakravarty

Supervision: Sanjeev Kumar Singh, Aparna Chakravarty

Validation: Sagar Mani Pradhan, Anandu Hari, Aparna Chakravarty, Kusum Rani, Vrinda Nampoothiri

Visualization: Sagar Mani Pradhan, Anandu Hari, Sanjeev Kumar Singh, Aparna Chakravarty

Writing-original draft: Sagar Mani Pradhan, Kusum Rani

Writing-review and editing: Sagar Mani Pradhan, Kusum Rani, Aparna Chakravarty, Vrinda

Nampoothiri, Anandu Hari, Fabia Edathadathil, Sanjeev Kumar Singh

## Acknowledgements

The authors would like to express their sincere gratitude to all individuals and organizations who contributed to the successful completion of this research. We are especially grateful to Durgga KN, Rio Varghese, Nalla Sanjay, Abin Joy, Niranjana Varma, Hemal Baghel, Ronak Sahu, Devanshu Bhardwaj, Aiswarya Sajeev, Praseedha Prasad Shetty, Arathi V, Chhaya Chauhan, Savitha A, Allied Health Sciences students, Amrita Vishwa Vidyapeetham, Faridabad, India for their time, cooperation, and valuable contributors as peer educators.

We also acknowledge the support of Wellcome Trust-funded programme CAMO-Net-India hub [grant ref: 226730/Z/22/Z) for facilitating and providing the necessary resources for conducting this study. Special appreciation is extended to colleagues and collaborators who assisted with various aspects of the project, including planning, implementation, data collection, and analysis. Finally, we thank our families and friends for their patience, understanding, and encouragement during the course of this research. The authors are grateful to all who contributed, directly or indirectly, to the completion of this work.

