## Supplemental Data for "Working with the future for the future: a peer-led educational intervention on antimicrobial resistance; a quasi-experimental study"

### SUPPLEMENTARY MATERIALS

S1. Table 1. Content of the module

| LESSON PLAN | OBJECTIVES | CONTENTS | REFERENCES FOR RESOURCES USED |
|---|---|---|---|
| **Lesson 01: Good Germs, Bad Germs** | • Familiarise the students with different classes of microorganisms.<br>• Help understand the differences between beneficial microorganisms and harmful microbes | • Introduction to microorganisms<br>• Classes of microorganisms<br>• Helpful and harmful microorganisms | https://youtu.be/jzjzqhfg6ec?si=gi2whkpxk0ihtwhy<br>https://youtu.be/vzpd009qtn4?si=j-pmszccciknilvw<br>https://sasuperbugs.org/ |
| **Lesson 02: Fightin** | • Define infection, immunity | **Section 1**<br>• Introduction to immunity | https://youtu.be/aq-f4rnuj3y?si=e-jfgfb7urutnxhq |

| | | | |
|---|---|---|---|
| **g The Bugs** | and antimicrobial agents<br>• Describe antibiotics<br>• Introduction to superbugs and antimicrobial resistance (AMR)<br>• Identify the causes and consequences of AMR.<br>• demonstrate how they can help prevent AMR in the country | • Tackling the microbes<br>• introduction to antimicrobial agents<br>**Section 2**<br>• Story about antibiotics<br>• Where does the antibiotic work?<br>• When antibiotics don't work!<br>• Introduction to superbugs- evolution and AMR<br>• cause and consequences of AMR | https://youtu.be/0zwjzcstd5m?si=nrw5mx11ifeaxde8<br>https://youtu.be/bgzj4bqbkmq?si=mnh51mpqm8nwdhba<br>https://sasuperbugs.org/<br>https://sasuperbugs.org/ |
| **Lesson 03: Bugs** | • Explain the importance of good | • Introduction to transmission of pathogens | https://youtu.be/iisgnbmfkvi?si=c3utawjbyg9no1q-resourcesrequired<br>https://youtu.be/6icsujnb_pm?si=0xfjur-m9nov4owk |

| | | | |
|---|---|---|---|
| **Won't Bug!** | hygiene practices.<br>• Demonstrate the correct steps of handwashing and hand sanitizing.<br>• Understand how infections spread through coughing, sneezing and touching.<br>• Practice respiratory etiquette to protect themselves and others | • Importance of handwashing<br>• Correct steps of handwashing and using hand sanitizers<br>• Respiratory etiquettes | https://youtu.be/iisgnbmfkvi?si=c3utawjbyg9no1q-resourcesrequired |

S2. Fig 1. Structured questionnaire used in the study

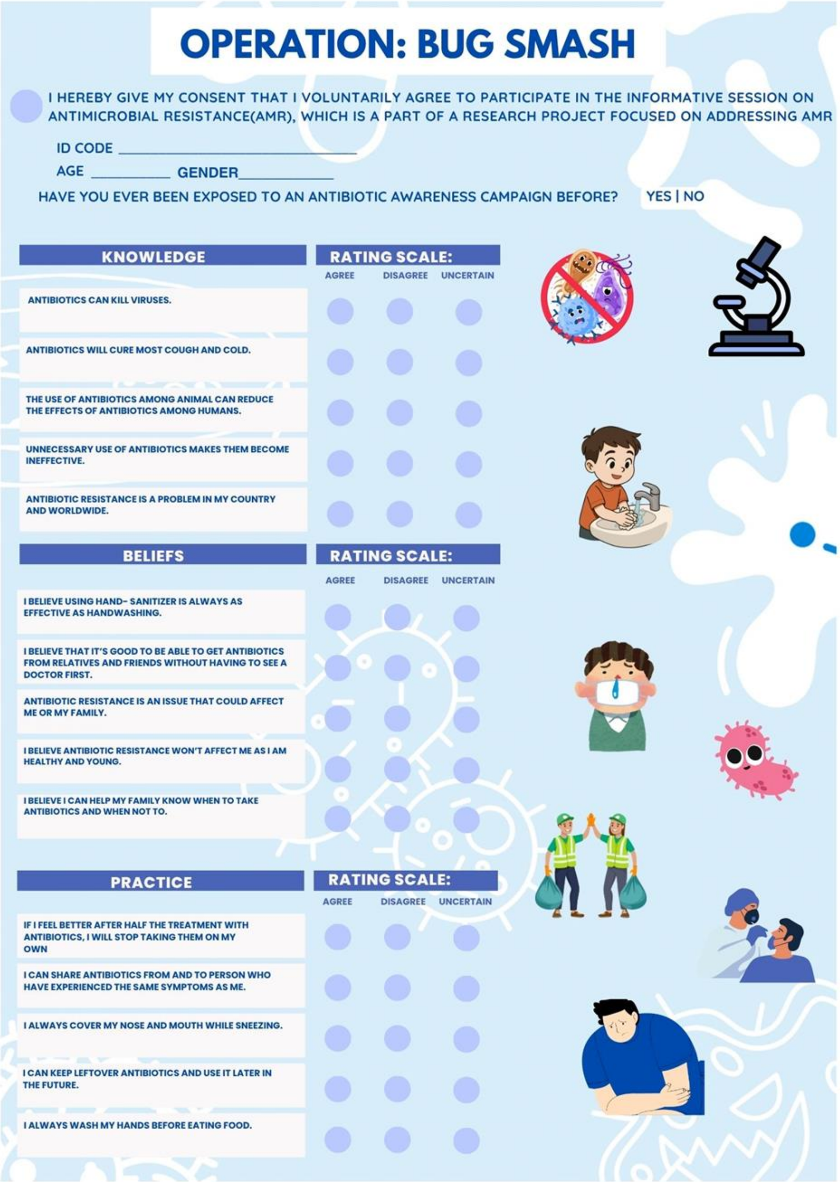

### OPERATION: BUG SMASH

I HEREBY GIVE MY CONSENT THAT I VOLUNTARILY AGREE TO PARTICIPATE IN THE INFORMATIVE SESSION ON ANTIMICROBIAL RESISTANCE(AMR), WHICH IS A PART OF A RESEARCH PROJECT FOCUSED ON ADDRESSING AMR

ID CODE ______________________________

AGE __________ GENDER____________

HAVE YOU EVER BEEN EXPOSED TO AN ANTIBIOTIC AWARENESS CAMPAIGN BEFORE? YES | NO

| KNOWLEDGE | AGREE | DISAGREE | UNCERTAIN |
|---|---|---|---|
| ANTIBIOTICS CAN KILL VIRUSES. | | | |
| ANTIBIOTICS WILL CURE MOST COUGH AND COLD. | | | |
| THE USE OF ANTIBIOTICS AMONG ANIMAL CAN REDUCE THE EFFECTS OF ANTIBIOTICS AMONG HUMANS. | | | |
| UNNECESSARY USE OF ANTIBIOTICS MAKES THEM BECOME INEFFECTIVE. | | | |
| ANTIBIOTIC RESISTANCE IS A PROBLEM IN MY COUNTRY AND WORLDWIDE. | | | |

| BELIEFS | AGREE | DISAGREE | UNCERTAIN |
|---|---|---|---|
| I BELIEVE USING HAND- SANITIZER IS ALWAYS AS EFFECTIVE AS HANDWASHING. | | | |
| I BELIEVE THAT IT'S GOOD TO BE ABLE TO GET ANTIBIOTICS FROM RELATIVES AND FRIENDS WITHOUT HAVING TO SEE A DOCTOR FIRST. | | | |
| ANTIBIOTIC RESISTANCE IS AN ISSUE THAT COULD AFFECT ME OR MY FAMILY. | | | |
| I BELIEVE ANTIBIOTIC RESISTANCE WON'T AFFECT ME AS I AM HEALTHY AND YOUNG. | | | |
| I BELIEVE I CAN HELP MY FAMILY KNOW WHEN TO TAKE ANTIBIOTICS AND WHEN NOT TO. | | | |

| PRACTICE | AGREE | DISAGREE | UNCERTAIN |
|---|---|---|---|
| IF I FEEL BETTER AFTER HALF THE TREATMENT WITH ANTIBIOTICS, I WILL STOP TAKING THEM ON MY OWN | | | |
| I CAN SHARE ANTIBIOTICS FROM AND TO PERSON WHO HAVE EXPERIENCED THE SAME SYMPTOMS AS ME. | | | |
| I ALWAYS COVER MY NOSE AND MOUTH WHILE SNEEZING. | | | |
| I CAN KEEP LEFTOVER ANTIBIOTICS AND USE IT LATER IN THE FUTURE. | | | |
| I ALWAYS WASH MY HANDS BEFORE EATING FOOD. | | | |

S3. Fig. 2. Information leaflet sent to the parents

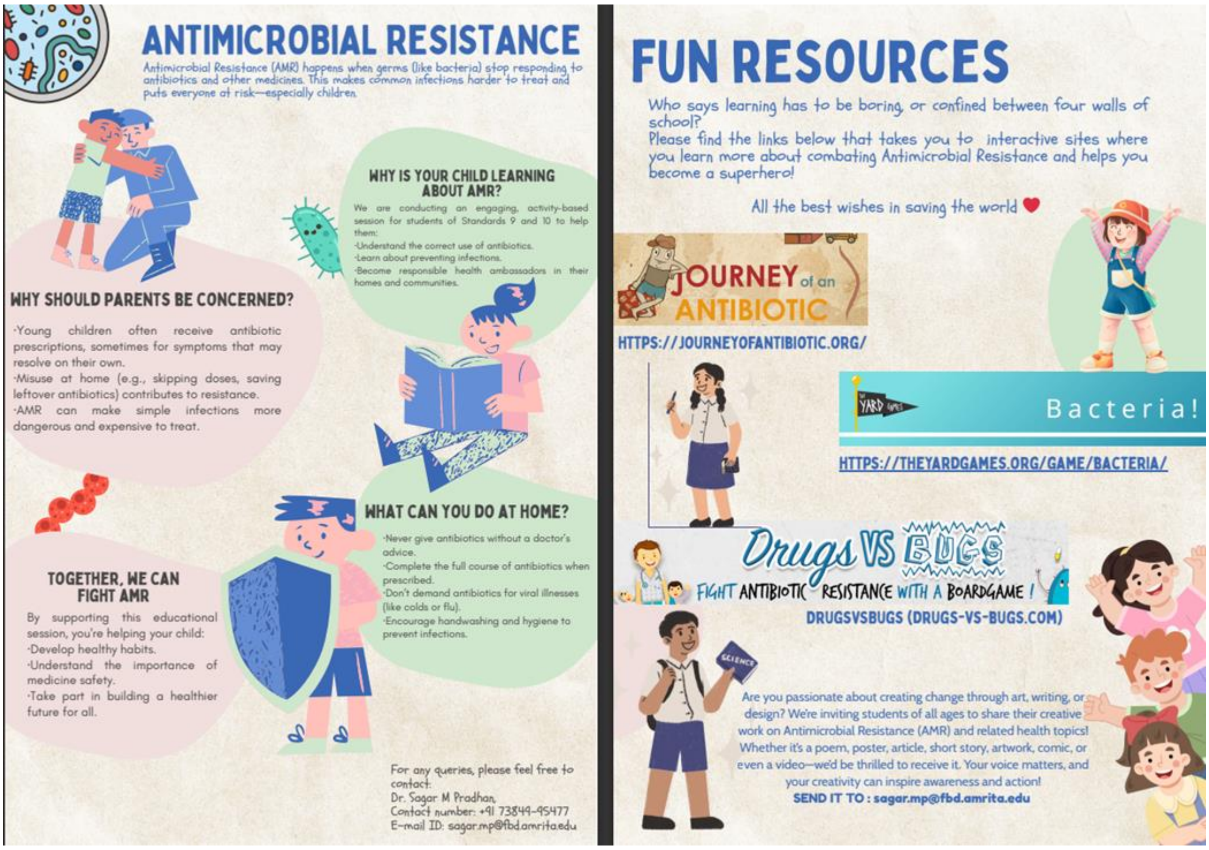


S4. Fig. 3. Art Reflective Worksheet

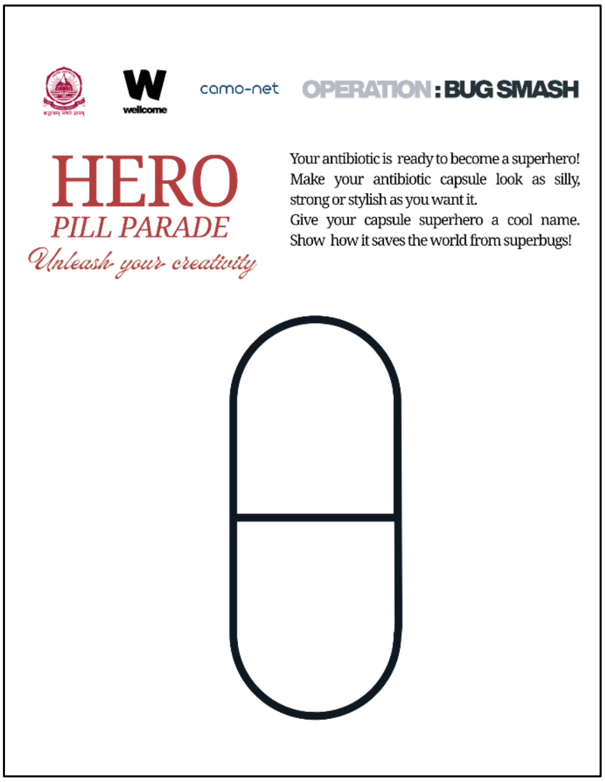

S5. Role-play script:

https://docs.google.com/document/d/1UxwPVnaPAEi_rap3uNkd6ZYsagMQyxpK/edit?usp=sharing&ouid=106997435465702419247&rtpof=true&sd=true
